## Supplementary Table 1 for "Respiratory cryptosporidiosis in Malawian children with diarrheal disease"

**Supplementary Table 1.** Full characteristics of study population at enrollment

| Characteristic | Children with diarrhea (n=162) | Cryptosporidium-positive (n=36) | | Cryptosporidium-negative (n=124) | P value |
| --- | --- | --- | --- | --- | --- |
| Number of household members (SD) | 4.5 (1.4) | 4.1 (1.2) | | 4.6 (1.4) | 0.061 |
| Adults ≥18 years | 2.1 (0.8) | 1.9 (0.5) | | 2.1 (0.8) | 0.072 |
| Children <18 years | 2.3 (1.1) | 2.1 (1.1) | | 2.3 (1.1) | 0.322 |
| Children <5 years | 1.2 (0.5) | 1.2 (0.4) | | 1.2 (0.6) | 0.107 |
| Number of people living/sleeping regularly in the compound for the past 6 months (SD) | 4.4 (1.4) | 4.1 (1.2) | | 4.5 (1.4) | 0.094 |
| Shared pit latrine/toilet for disposable of feces (%) |  |  | |  |  |
| 2 households | 114 (70%) | 27 (75%) | | 84 (69%) | 0.442 |
| 3-5 households | 45 (28%) | 8 (22%) | | 37 (30%) |  |
| ≥6 households | 3 (2%) | 1 (3%) | | 1 (1%) |  |
| Residential animals in the compound (%) | 107 (66%) | 21 (20%) | | 84 (68%) | 0.359 |
| Goat | 6/107 (6%) | 2 (5%) | | 4 (5%) | 0.345 |
| Cow | 0 | - | | - | - |
| Pig | 2 (2%) | 1 (5%) | | 1 (1%) | 0.362 |
| Fowl | 31 (29%) | 7 (33%) | | 24 (29%) | 0.669 |
| Dog | 31 (29%) | 4 (19%) | | 26 (31%) | 0.280 |
| Cat | 17 (16%) | 5 (24%) | | 11 (13%) | 0.222 |
| Rodents | 72 (67%) | 13 (62%) | | 59 (70%) | 0.462 |
| **Water source** |  |  | |  |  |
| Drinking (%) |  |  |  | |  |
| Piped water | 126 (78%) | 28 (80%) | 95 (77%) | | 0.728 |
| Well/borehole | 36 (22%) | 7 (20%) | | 28 (23%) |  |
| Pond/lake, river/stream | 0 | 0 | | 0 |  |
| Cooking (%) |  |  |  | |  |
| Piped water | 122 (75%) | 28 (78%) | 91 (75%) | | 0.776 |
| Well/borehole | 40 (25%) | 8 (22%) | 31 (25%) | |  |
| Pond/lake, river/stream | 0 | 0 | | 0 |  |
| Bathing water (%) |  |  |  | |  |
| Piped water | 101 (62%) | 23 (64%) | 75 (62%) | | 0.508 |
| Well/borehole | 53 (33%) | 10 (28%) | | 42 (34%) |  |
| Pond/lake, river/stream | 8 (5%) | 3 (8%) | | 5 (5%) |  |
| Utensil water (%) |  |  | |  |  |
| Piped water | 112 (69%) | 23 (66%) | | 86 (70%) | 0.073 |
| Well/borehole | 48 (30%) | 10 (29%) | | 37 (30%) |  |
| Pond/lake, river/stream | 2 (1%) | 2 (6%) | | 0 |  |
| Washing water (%) |  |  |  | |  |
| Piped water | 82 (50%) | 16 (46%) | 65 (53%) | | 0.447 |
| Well/borehole | 41 (25%) | 8 (23%) | | 32 (26%) |  |
| Pond/lake, river/stream | 39 (24%) | 11 (31%) | | 26 (21%) |  |
| Time taken to get to water and return, minutes^a^ (SD) | 26.9 (14.1) | 26 (13.8) | | 27.1 (14.1) | 0.708 |
| Number of trips made to fetch water per week (SD) | 27.1 (10) | 25 (7.9) | | 27.7 (10.4) | 0.192 |
| Treated water for drinking^b^ (%) | 23 (14%) | 6 (17%) | | 17 (14%) | 0.596 |
| Child given untreated drinking water in the last week before admission (%) | 74 (46%) | 15 (43%) | | 57 (46%) | 0.715 |

^a^Boiling, or using cloth chlorine, ceramic or other filters.

^b^Unable to assess sputum quality due to poor appearance.
